# Tumor-Specific Divergence of Tumor-Associated Macrophage Prognostic Effects Across TCGA Lung and Melanoma Cohorts

**DOI:** 10.64898/2026.02.23.26346900

**Authors:** Steven Lehrer, Peter H. Rheinstein

## Abstract

**Background:** Tumor-associated macrophages (TAMs) display context-dependent functional polarization, but whether their prognostic impact is consistent across tumor types remains unclear.

**Methods:** We analyzed RNA-sequencing and clinical data from The Cancer Genome Atlas (TCGA) lung adenocarcinoma (LUAD; n=648), lung squamous carcinoma (LUSC; n=623), and melanoma (SKCM; n=466). Cox proportional hazards models adjusted for age and AJCC stage evaluated per–standard deviation (SD) expression of TAM markers (FOLR2, TREM2) and T-cell markers (CD8A, CXCL9). Cross-histology interaction terms tested divergence between LUAD and LUSC.

**Results:** In melanoma, higher FOLR2 (HR 0.87), TREM2 (HR 0.83), CD8A (HR 0.69), and CXCL9 (HR 0.67) independently predicted improved survival. LUAD showed largely neutral macrophage effects. In contrast, LUSC demonstrated an adverse association for FOLR2 (HR 1.28). Interaction analysis confirmed significant divergence for FOLR2 and TREM2 between LUAD and LUSC.

**Conclusions:** TAM-associated prognostic effects reverse by tumor histology, supporting tumor-context–dependent macrophage polarization and informing macrophage-targeted therapeutic strategies.

## Introduction

The tumor microenvironment (TME) critically shapes oncologic outcomes [1]. Among immune populations, tumor-associated macrophages (TAMs) are highly plastic and exhibit context-dependent phenotypes ranging from pro-inflammatory to immunosuppressive states. Although TAM infiltration has been variably associated with prognosis across tumor types, systematic cross-histology comparisons remain limited [2].

Mateus-Tique et al. describe a strategy to overcome immune suppression in solid tumors by targeting tumor-associated macrophages (TAMs) with “armored” CAR-T cells engineered to secrete IL-12 [3]. The authors focus on TAM subsets characterized by expression of FOLR2 or TREM2, markers enriched in immunosuppressive macrophages across multiple solid tumors. Rather than targeting cancer cells directly, they redirect CAR-T cells against these myeloid populations to remodel the tumor microenvironment (TME). Conventional anti-FOLR2 CAR-T cells effectively trafficked to tumors and selectively depleted FOLR2+ macrophages but produced only modest survival benefit. Armoring these CAR-T cells with IL-12 dramatically enhanced therapeutic efficacy. High-dose IL-12 CAR-T was toxic; however, low-dose IL-12– expressing anti-TAM CAR-T achieved durable tumor regression without lymphodepletion and without overt systemic toxicity. In aggressive metastatic ovarian and lung cancer models, treatment reduced tumor burden, extended survival, and in some cases produced complete remission.

Mechanistically, spatial transcriptomics demonstrated that IL-12 anti-TAM CAR-T cells did more than deplete suppressive macrophages. They induced profound reprogramming of the TME. Folr2^high immunosuppressive macrophages were nearly eliminated and replaced by CXCL9+ and IL-1β+ macrophage subsets with immunostimulatory, antigen-presenting phenotypes. The ratio of stimulatory to suppressive macrophages increased dramatically, and this reprogrammed state persisted even after CAR-T contraction, indicating durable TME remodeling.Concomitantly, there was a marked expansion of endogenous CD8+ T cells, reduction of regulatory T cells, increased granzyme B and IFN-γ expression, and formation of memory-like T cell populations. Importantly, tumor control depended in part on FAS upregulation on cancer cells, establishing an IL-12–FAS axis whereby endogenous CD8+ T cells mediated FAS-dependent tumor cell killing. Similar effects were observed with IL-12–armored anti-TREM2 CAR-T cells in metastatic lung cancer models, supporting TREM2 as an alternative TAM target [3].

Overall, the study establishes that myeloid-directed, IL-12-armored CAR-T cells can reset the tumor immune ecosystem, converting “cold” tumors into “hot” tumors by eliminating suppressive TAMs, inducing CXCL9+ macrophages, expanding endogenous tumor-reactive CD8+ T cells, and driving durable anti-tumor immunity [3]. The work reframes macrophages not simply as passive suppressors but as programmable gatekeepers of anti-tumor immunity, and positions TAM-targeted IL-12 CAR-T therapy as a broadly applicable strategy for solid cancers.

Melanoma represents an immune-responsive malignancy characterized by high tumor mutational burden (TMB) and sensitivity to checkpoint blockade [4]. In contrast, lung cancers exhibit heterogeneous immune landscapes, with lung squamous carcinoma (LUSC) often demonstrating distinct stromal and macrophage features compared to lung adenocarcinoma (LUAD) [5].

We hypothesized that TAM-associated gene expression may exert tumor-type–specific prognostic effects. To test this, we performed a comparative analysis of TAM and T-cell markers across The Cancer Genome Atlas (TCGA) LUAD (lung adenocarcinoma), LUSC (lung squamous carcinoma), and SKCM (melanoma) cohorts.

## Methods

### Data Sources

Bulk RNA-seq and clinical data were obtained from The Cancer Genome Atlas (TCGA) with the UCSC Xena Browser [6] from the following data sets: LUAD, LUSC, SKCM.

Gene Selection Markers analyzed: TAM-associated: FOLR2, TREM2 [7]; T-cell/interferon-gamma (IFNγ)-associated: CD8A, CXCL9 [8]

Expression values were log-transformed and standardized (per standard deviation SD). Survival analysis primary endpoint was overall survival. Cox proportional hazards models were adjusted for Age at diagnosis and AJCC pathologic stage.

Models: Per-Standard Deviation (SD) gene models, TAM × CD8 interaction models, Cross-histology interaction models, and 4-group Kaplan–Meier (median split FOLR2 and CD8A)

## Results

### Total analyzed samples are tabulated in table 1

Figure 1 shows Per–standard deviation Cox models of TAM and T-cell markers across TCGA lung and melanoma cohorts. Forest plots show hazard ratios (HR) for overall survival per one–standard deviation (SD) increase in gene expression for FOLR2, TREM2 (tumor-associated macrophage markers), and CD8A, CXCL9 (T-cell/IFNγ-associated markers). Cox proportional hazards models were adjusted for age at diagnosis and AJCC pathologic stage. Vertical dashed line indicates HR = 1.0. In melanoma (SKCM), all four markers were independently associated with improved survival (HR < 1). In lung adenocarcinoma (LUAD), effects were largely neutral. In lung squamous carcinoma (LUSC), FOLR2 demonstrated an adverse association (HR > 1), indicating tumor-type–specific divergence of macrophage-associated prognostic effects.

**Table 1.**
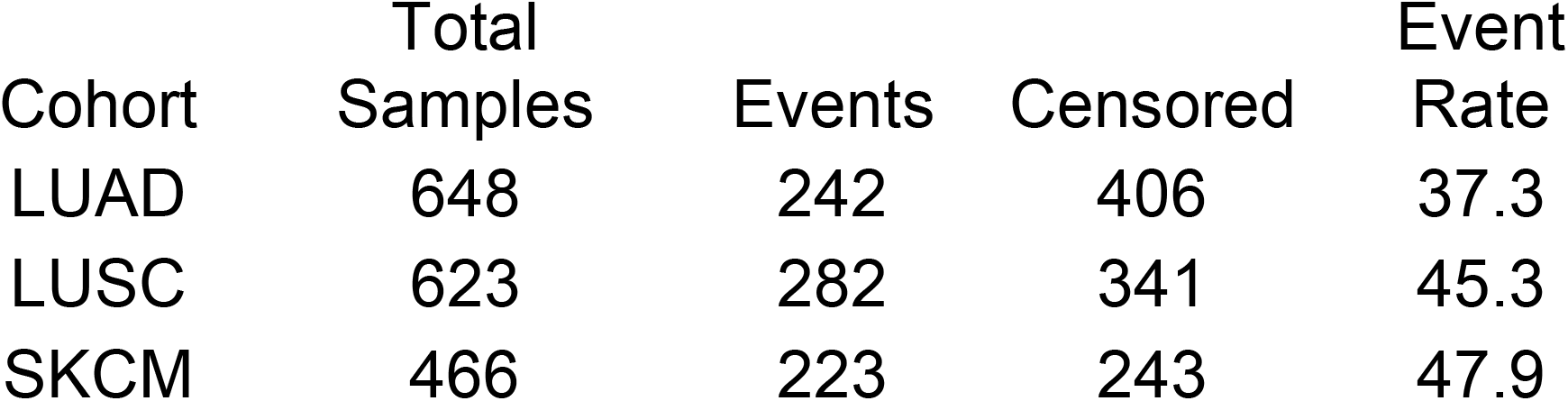
Total analyzed samples.

| Cohort | Total<br>Samples | Events | Censored | Event<br>Rate |
| --- | --- | --- | --- | --- |
| LUAD | 648 | 242 | 406 | 37.3 |
| LUSC | 623 | 282 | 341 | 45.3 |
| SKCM | 466 | 223 | 243 | 47.9 |

**Figure 1.**
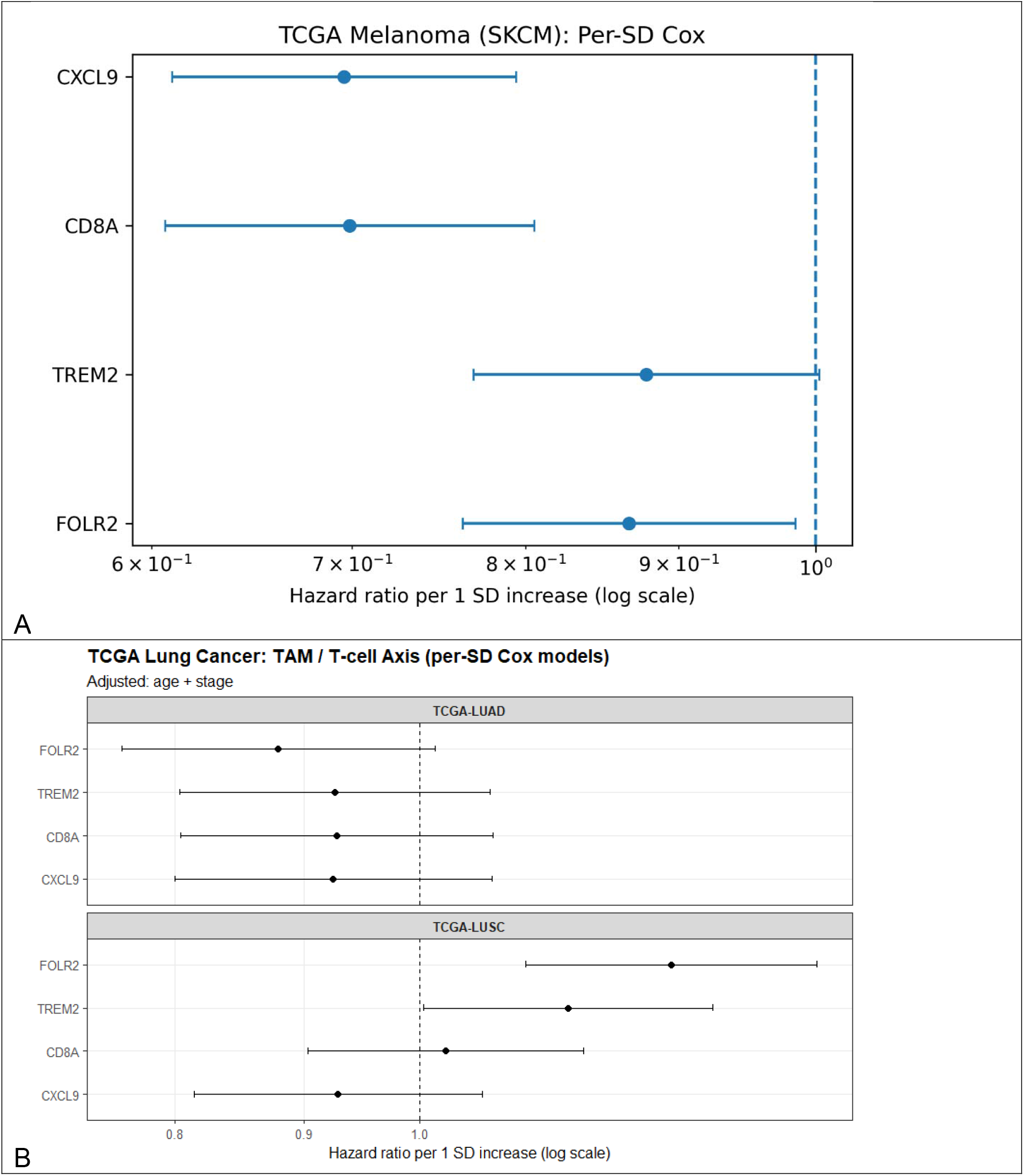
Per–standard deviation Cox models of TAM and T-cell markers across TCGA lung and melanoma cohorts. Forest plots show hazard ratios (HR) for overall survival per one–standard deviation (SD) increase in gene expression for FOLR2, TREM2 (tumor-associated macrophage markers), and CD8A, CXCL9 (T-cell/IFNγ-associated markers). Cox proportional hazards models were adjusted for age at diagnosis and AJCC pathologic stage. Vertical dashed line indicates HR = 1.0. (A) In melanoma (SKCM), all four markers were independently associated with improved survival (HR < 1). (B) In lung adenocarcinoma (LUAD), effects were largely neutral. In lung squamous carcinoma (LUSC), FOLR2 demonstrated an adverse association (HR > 1), indicating tumor-type–specific divergence of macrophage-associated prognostic effects. Error bars represent 95% confidence intervals.

Figure 2 shows Kaplan–Meier survival analysis of FOLR2 and CD8A stratified immune groups in melanoma. FOLR2 is being used as a marker of Tumor Associated Macrophages (TAMs). Kaplan–Meier curves for overall survival in TCGA-SKCM were stratified by median expression of FOLR2 (TAM marker) and CD8A (T-cell marker), generating four groups: TAM_high/CD8_high, TAM_high/CD8_low, TAM_low/CD8_high, and TAM_low/CD8_low. High CD8A expression was associated with improved survival, and high FOLR2 expression tracked with favorable outcomes in melanoma, consistent with an immune-inflamed tumor phenotype.

**Figure 2.**
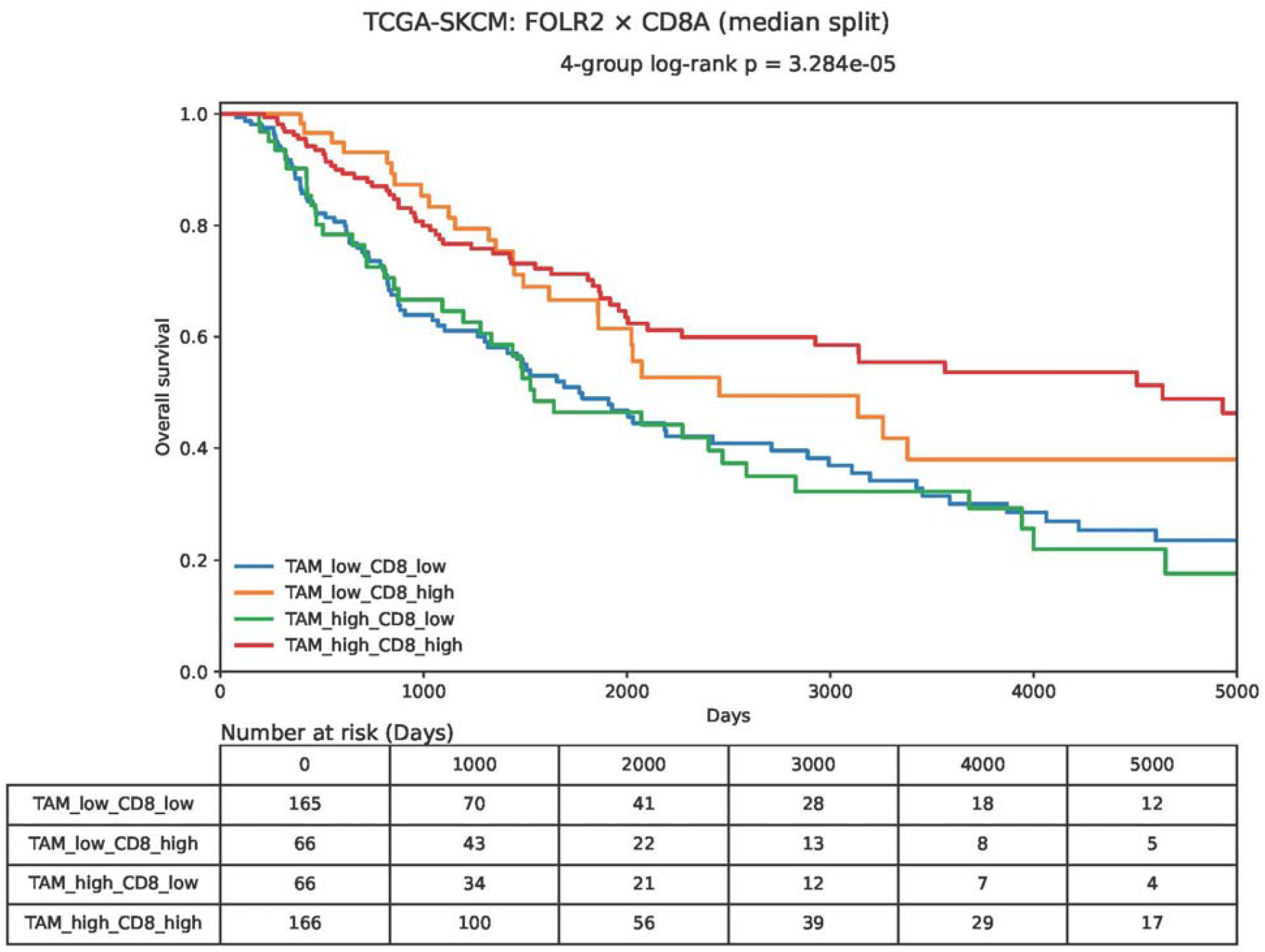
Kaplan–Meier survival analysis of FOLR2 and CD8A stratified immune groups in melanoma. FOLR2 is being used as a marker of Tumor Associated Macrophages (TAMs). Kaplan–Meier curves for overall survival in TCGA-SKCM stratified by median expression of FOLR2 (TAM marker) and CD8A (T-cell marker), generating four groups: TAM_high/CD8_high, TAM_high/CD8_low, TAM_low/CD8_high, and TAM_low/CD8_low. Log-rank p-value is shown on the plot. Number-at-risk table is displayed below the survival curves at prespecified time intervals. High CD8A expression was associated with improved survival, and high FOLR2 expression tracked with favorable outcomes in melanoma, consistent with an immune-inflamed tumor phenotype.

Figure 3 shows Kaplan–Meier survival analysis of FOLR2 and CD8A stratified immune groups in lung squamous carcinoma (LUSC). Kaplan–Meier curves for TCGA-LUSC were stratified by median FOLR2 and CD8A expression. Four immune groups were generated as in Figure 2. In contrast to melanoma, elevated FOLR2 expression was associated with worse overall survival in LUSC.

**Figure 3.**
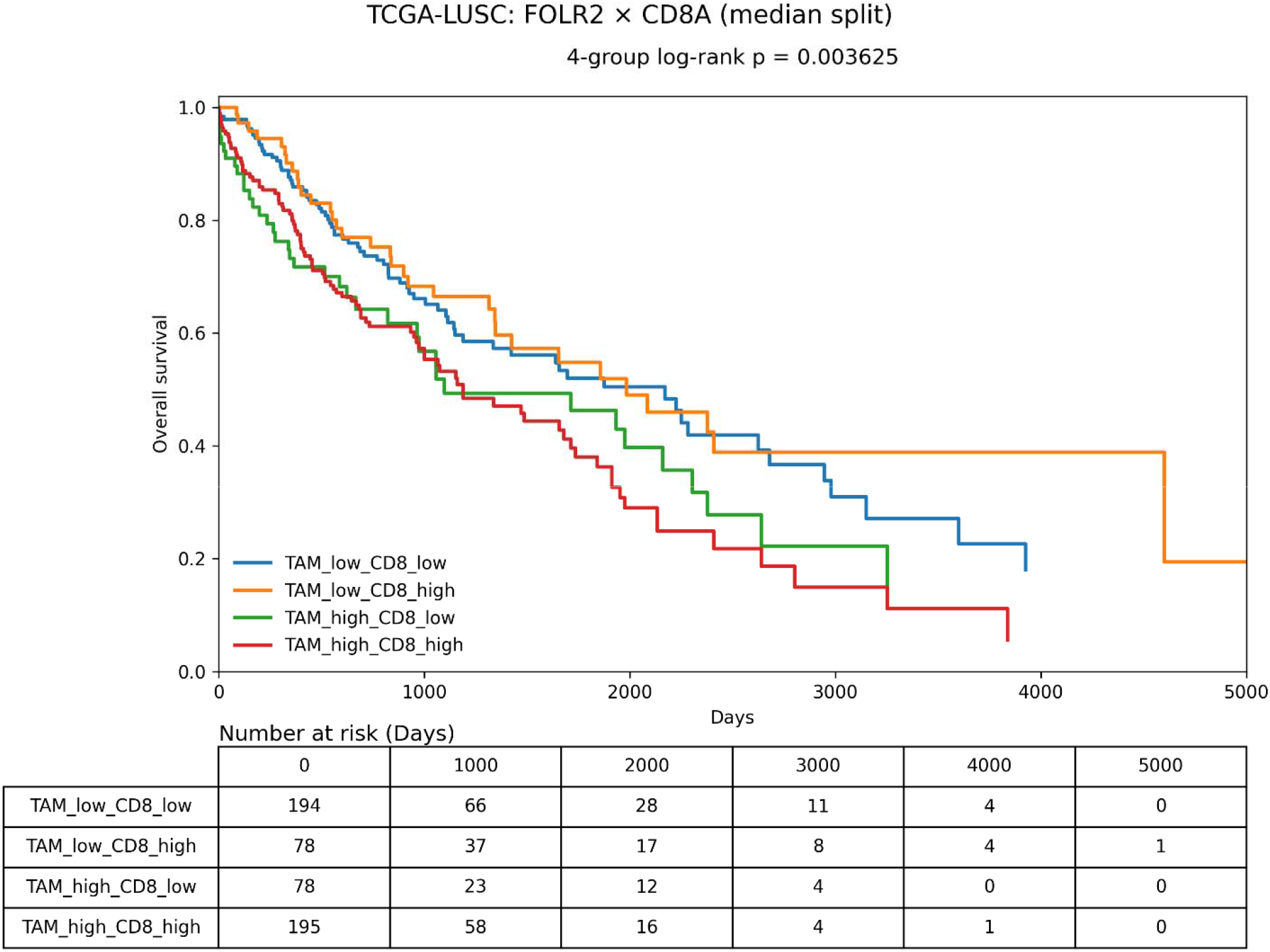
Kaplan–Meier survival analysis of FOLR2 and CD8A stratified immune groups in lung squamous carcinoma (LUSC). Kaplan–Meier curves for TCGA-LUSC stratified by median FOLR2 and CD8A expression. Four immune groups were generated as in Figure 2. In contrast to melanoma, elevated FOLR2 expression was associated with worse overall survival in LUSC. Log-rank p-value indicates significant survival divergence across immune strata. Number-at-risk table is shown below the plot. These results demonstrate reversal of macrophage-associated prognostic direction compared to melanoma.

Figure 4 shows cross-histology interaction analysis of TAM markers between LUAD and LUSC. Forest plot illustrates hazard ratios for per-SD gene expression in LUAD (reference) and multiplicative interaction terms for LUSC versus LUAD. Cox models included gene expression, histology indicator, and gene × histology interaction term, adjusted for age and stage. Significant interaction was observed for FOLR2 (p < 0.001) and TREM2 (p = 0.03), confirming histology-specific divergence in macrophage-associated prognostic effects. CD8A and CXCL9 did not demonstrate significant interaction.

**Figure 4.**
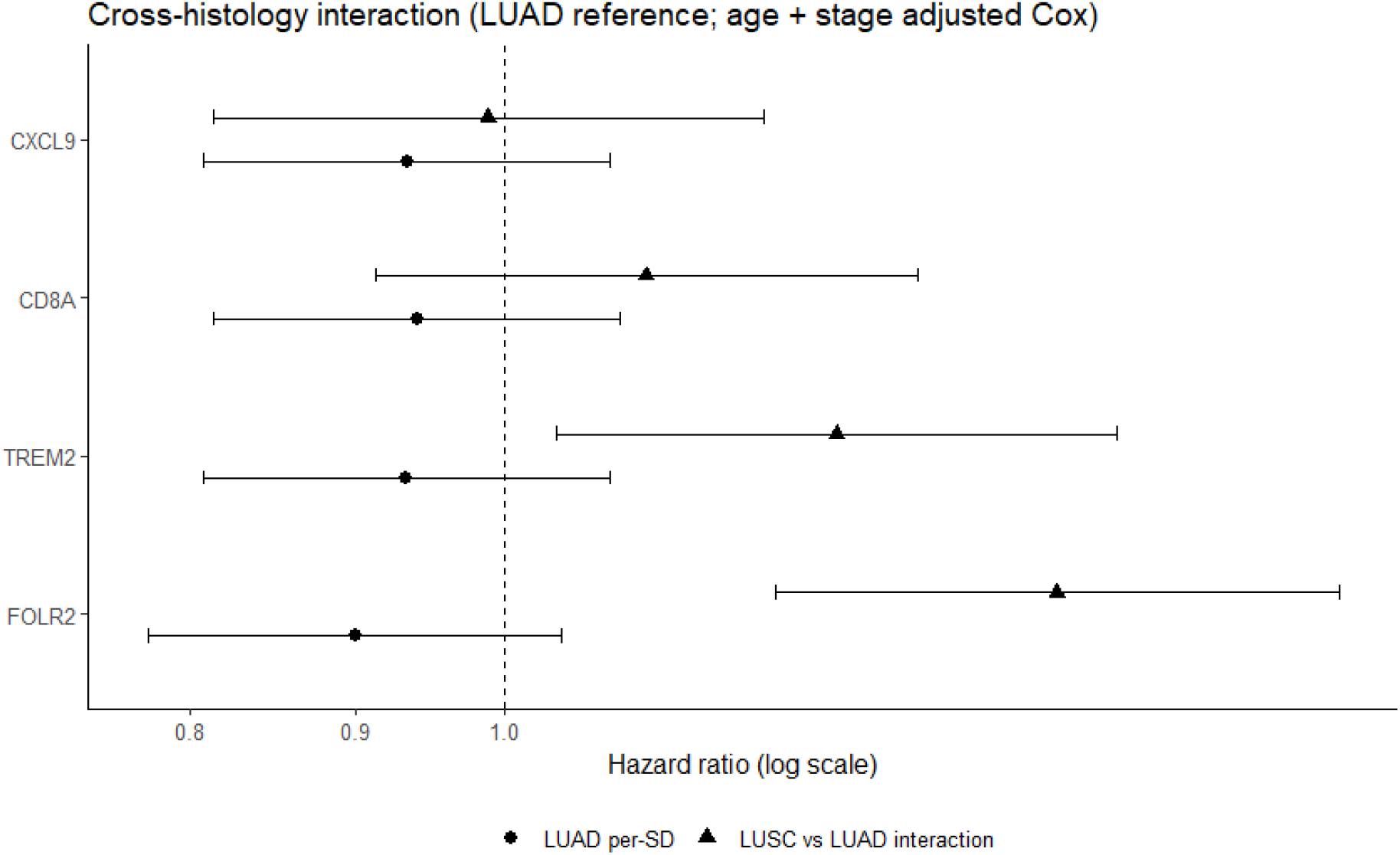
Cross-histology interaction analysis of TAM markers between LUAD and LUSC. Forest plot showing hazard ratios for per-SD gene expression in LUAD (reference) and multiplicative interaction terms for LUSC versus LUAD. Cox models included gene expression, histology indicator, and gene × histology interaction term, adjusted for age and stage. Significant interaction was observed for FOLR2 (p < 0.001) and TREM2 (p = 0.03), confirming histology-specific divergence in macrophage-associated prognostic effects. CD8A and CXCL9 did not demonstrate significant interaction.

Figure 5 is a summary model of tumor-type–specific macrophage polarization, a conceptual schematic illustrating differential TAM functional states across tumor types. In melanoma, elevated FOLR2 and TREM2 expression correlate with CD8A/CXCL9-associated immune activation and improved survival. In lung squamous carcinoma, elevated FOLR2 expression correlates with adverse outcomes, consistent with immunosuppressive macrophage polarization. LUAD exhibits intermediate or neutral effects. This model proposes tumor-context–dependent macrophage functional polarization influencing clinical outcomes.

**Figure 5.**
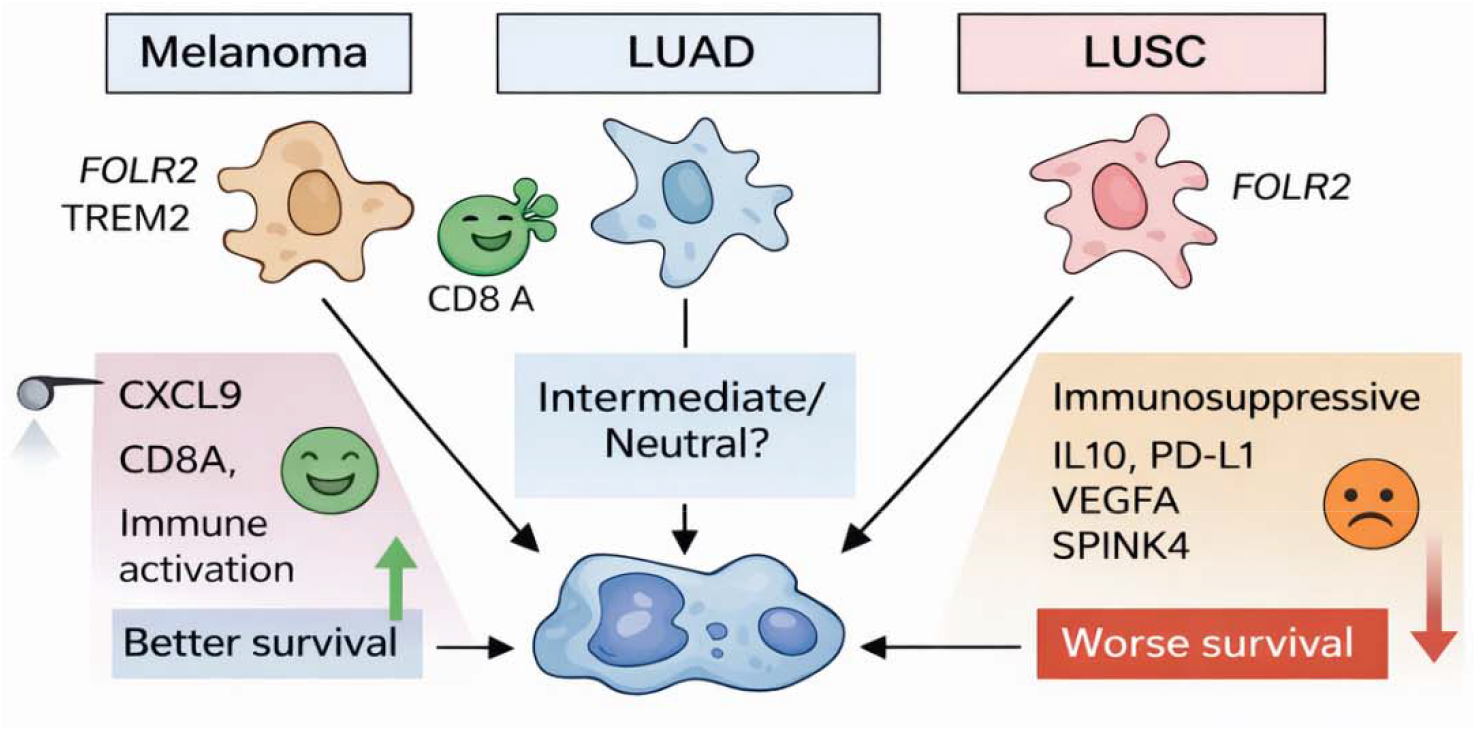
Summary model of tumor-type–specific macrophage polarization. Conceptual schematic illustrating differential TAM functional states across tumor types. In melanoma, elevated FOLR2 and TREM2 expression correlate with CD8A/CXCL9-associated immune activation and improved survival. In lung squamous carcinoma, elevated FOLR2 expression correlates with adverse outcomes, consistent with immunosuppressive macrophage polarization. LUAD exhibits intermediate or neutral effects. This model proposes tumor-context–dependent macrophage functional polarization influencing clinical outcomes.

## Discussion

Tumor-associated macrophages (TAMs) are increasingly recognized not merely as passive components of the tumor microenvironment (TME), but as dynamic regulators of immune suppression or immune activation depending on context [9]. In their recent Cancer Cell study, Mateus-Tique et al. demonstrated that FOLR2 and TREM2 TAMs can be therapeutically targeted with IL-12–armored CAR-T cells to deplete immunosuppressive macrophage populations, induce CXCL9 immunostimulatory macrophages, expand endogenous CD8 T cells, and achieve durable tumor regression in metastatic ovarian and lung cancer models [3]. Their work reframes macrophages as programmable gatekeepers of anti-tumor immunity and establishes an IL-12–driven macrophage–T cell axis as a central mechanism of TME remodeling.

Our cross-tumor analysis of TCGA lung adenocarcinoma (LUAD), lung squamous carcinoma (LUSC), and melanoma (SKCM) provides complementary human evidence supporting this framework. Notably, we observed that the prognostic impact of TAM-associated gene expression diverges sharply by tumor histology. In melanoma, higher expression of FOLR2 and TREM2 was independently associated with improved overall survival and coincided with strong CD8A and CXCL9 signals, consistent with an immune-inflamed phenotype. In contrast, in LUSC, FOLR2 expression was associated with worse survival, whereas LUAD demonstrated largely neutral effects. These findings suggest that TAM-associated gene expression does not uniformly signify immunosuppression but instead reflects tumor-type–specific macrophage polarization states.

The mechanistic insights from Mateus-Tique et al. help contextualize these observations [3]. In preclinical models, IL-12–armored anti-TAM CAR-T cells eliminated FOLR2 high immunosuppressive macrophages and drove expansion of CXCL9 and IL-1β macrophage subsets enriched for antigen presentation and T-cell recruitment. This macrophage reprogramming established a self-sustaining IL-12/IFNγ circuit that expanded endogenous cytotoxic CD8 T cells and mediated FAS-dependent tumor cell killing.

The Fas-dependent tumor cell killing pathway is an extrinsic apoptosis mechanism where Fas ligand (FasL) on immune cells (CTLs, NK cells) binds to the Fas receptor (CD95) on tumor cells. This triggers trimerization, recruiting FADD and caspase-8/10 to form the death-inducing signaling complex (DISC), which activates caspases-3/7 to induce apoptosis [10].

Importantly, the balance between immunosuppressive FOLR2 high macrophages and CXCL9 immunostimulatory macrophages in the Mateus-Tique et al. study determined therapeutic outcome [3].

Our human data suggest that this balance may differ intrinsically across tumor types. In melanoma, where high tumor mutational burden and interferon signaling are hallmarks of immune responsiveness, elevated FOLR2 and TREM2 expression may reflect macrophage populations embedded within an IFNγ-rich, CXCL9-driven T-cell–inflamed environment [8]. In this setting, macrophage abundance correlates with effective immune surveillance and improved survival. Conversely, in LUSC, FOLR2 expression may represent expansion of immunosuppressive TAM subsets lacking the accompanying IFNγ/CXCL9 activation program necessary to support productive T-cell immunity. The neutral pattern observed in LUAD suggests a more heterogeneous macrophage landscape in which suppressive and stimulatory states may coexist without a dominant prognostic signal.

Importantly, our interaction analyses demonstrate that these differences are not simply quantitative but represent statistically significant histology-specific divergence in macrophage-associated hazard ratios. Thus, macrophage biology appears to be context-dependent at the tumor-type level. This observation has direct translational implications. The Mateus-Tique study shows that forced IL-12 delivery can convert suppressive TAMs into CXCL9 immunostimulatory macrophages and restore anti-tumor immunity [3]. Our findings imply that the need for such reprogramming may vary across cancers. Tumors in which TAM-associated gene expression already tracks with favorable prognosis (e.g., melanoma) may contain macrophage populations that are partially aligned with anti-tumor immunity, whereas tumors in which TAM markers predict worse survival (e.g., LUSC) may require more aggressive macrophage-directed reprogramming strategies.

These data also underscore a broader conceptual point: TAM marker expression alone cannot be interpreted as uniformly pro- or anti-tumoral without accounting for the surrounding immune context. Bulk RNA-based macrophage signatures likely capture mixtures of suppressive FOLR2 high populations and CXCL9 immunostimulatory subsets, with the relative dominance of each varying by tumor type. This model aligns closely with the macrophage plasticity and state transitions documented by Mateus-Tique et al., in which depletion of suppressive macrophages and IL-12–mediated T-cell activation shift the TME toward a CXCL9 macrophage–CD8 T cell feedback loop.

Several limitations warrant consideration. Our study relies on retrospective TCGA bulk RNA data and cannot resolve macrophage subpopulations at single-cell resolution. Bulk RNA does not identify macrophage subtypes. Independent melanoma RNA validation in a second data set would be worthwhile.

Nonetheless, the consistency of tumor-type–specific divergence across multiple markers and the concordance with mechanistic macrophage reprogramming data from preclinical models strengthen the biological plausibility of our conclusions. Future work integrating single-cell transcriptomics with survival outcomes across tumor types will further clarify the macrophage state compositions underlying these divergent prognostic effects.

In summary, while previous literature has variably associated TAMs with both good and bad outcomes, our study explicitly quantifies the statistical interaction between histology and gene expression to confirm that these differences are significant, not just qualitative. The most significant finding is the reversal of hazard ratios for the same macrophage markers across different histologies. Melanoma (SKCM) TAM markers (FOLR2, TREM2) are protective, predicting improved survival. In Lung Squamous Carcinoma (LUSC) the same markers (specifically FOLR2) are adverse, predicting worse survival. In Lung Adenocarcinoma (LUAD) the markers show a neutral or non-significant effect.

Our cross-histology survival analysis provides human correlative validation of the macrophage reprogramming paradigm established by Mateus-Tique et al. [3]. We demonstrate that the prognostic meaning of TAM-associated gene expression is tumor-type–specific and likely reflects the balance between immunosuppressive and immunostimulatory macrophage states. These findings reinforce the translational relevance of macrophage-targeted immunotherapies and highlight the importance of tumor-context–aware strategies when developing TAM-directed interventions.

Clinical Implications are multiple. Macrophage depletion strategies may have divergent effects across tumor types. Histology-specific biomarker stratification may be necessary. Companion diagnostic approaches should incorporate tumor context. Our findings highlight the tumor-type specificity of macrophage biology and caution against generalized macrophage-targeting strategies.

## Conclusion

TAM prognostic impact diverges sharply by tumor histology, being protective in melanoma and adverse in lung squamous carcinoma. These results underscore the need for tumor-specific macrophage-targeted therapeutic strategies.

## Data Availability

All data analyzed in this study are publicly available. RNA sequencing expression data and corresponding clinical annotations for TCGA lung adenocarcinoma (LUAD), lung squamous cell carcinoma (LUSC), and skin cutaneous melanoma (SKCM) were obtained from The Cancer Genome Atlas (TCGA) via the Genomic Data Commons (GDC) Data Portal (https://portal.gdc.cancer.gov/). Processed gene expression matrices were downloaded from the UCSC Xena browser (https://xena.ucsc.edu/), which provides harmonized TCGA datasets. Clinical survival data were obtained from the GDC clinical data files associated with each tumor type. No controlled-access data were used. All analyses were performed on de-identified, publicly available datasets.

https://xenabrowser.net/

## References

[1] Wang Q, Shao X, Zhang Y, Zhu M, Wang FXC, Mu J, Li J, Yao H, Chen K (2023) Role of tumor microenvironment in cancer progression and therapeutic strategy. Cancer Med 12, 11149–11165.

[2] Saeed AF (2025) Tumor-Associated Macrophages: Polarization, Immunoregulation, and Immunotherapy. Cells 14.

[3] Mateus-Tique J, Lakshmi A, Singh B, Iyer R, Sánchez-Paulete AR, Falcomatà C, Lin M, Pantsulaia G, Tepper A, Nguyen T, Amabile A, Mollaoglu G, Pia L, Chhamalwan D, Le Berichel J, Potak H, Colonna M, Baccarini A, Brody J, Merad M, Brown BD (2026) Armored macrophage-targeted CAR-T cells reset and reprogram the tumor microenvironment and control metastatic cancer growth. Cancer Cell.

[4] Timis T, Buruiana S, Dima D, Nistor M, Muresan XM, Cenariu D, Tigu AB, Tomuleasa C (2025) Advances in Cell and Immune Therapies for Melanoma. Biomedicines 13.

[5] Wang C, Yu Q, Song T, Wang Z, Song L, Yang Y, Shao J, Li J, Ni Y, Chao N, Zhang L, Li W (2022) The heterogeneous immune landscape between lung adenocarcinoma and squamous carcinoma revealed by single-cell RNA sequencing. Signal Transduction and Targeted Therapy 7, 289.

[6] Goldman M, Craft B, Swatloski T, Cline M, Morozova O, Diekhans M, Haussler D, Zhu J (2015) The UCSC Cancer Genomics Browser: update 2015. Nucleic Acids Res 43, D812–817.

[7] Nakamura K, Smyth MJ (2020) TREM2 marks tumor-associated macrophages. Signal Transduction and Targeted Therapy 5, 233.

[8] Nalio Ramos R, Missolo-Koussou Y, Gerber-Ferder Y, Bromley CP, Bugatti M, Núñez NG, Tosello Boari J, Richer W, Menger L, Denizeau J, Sedlik C, Caudana P, Kotsias F, Niborski LL, Viel S, Bohec M, Lameiras S, Baulande S, Lesage L, Nicolas A, Meseure D, Vincent-Salomon A, Reyal F, Dutertre C-A, Ginhoux F, Vimeux L, Donnadieu E, Buttard B, Galon J, Zelenay S, Vermi W, Guermonprez P, Piaggio E, Helft J (2022) Tissue-resident FOLR2+ macrophages associate with CD8+ T cell infiltration in human breast cancer. Cell 185, 1189–1207.e1125.

[9] Bied M, Ho WW, Ginhoux F, Blériot C (2023) Roles of macrophages in tumor development: a spatiotemporal perspective. Cellular & Molecular Immunology 20, 983–992.

[10] Hu L, Lu J, Fan H, Niu C, Han Y, Caiyin Q, Wu H, Qiao J (2025) FAS mediates apoptosis, inflammation, and treatment of pathogen infection. Frontiers in Cellular and Infection Microbiology Volume 15-2025.

